# Disparities in Opioid-related Mortality Across United States Census Regions from 1999-2020

**DOI:** 10.1101/2022.02.10.22270803

**Authors:** Supriyanka Addimulam, Swapnil Gupta, Sindhuja Mahalingam, Namrata Walia

## Abstract

**Background:** Opioid-related mortality has been on a sharp rise in the decade. This study aims to provide insight into the difference in mortality between white and black population in various census regions of the United States between 1999-2020.

**Methods:** The data was extracted from multiple cause of death files from CDC Wonder database. The International Classification of Disease (ICD-10) codes used to extract data include F11 (mental and behavioral disorders due to use of opioids); T40.0 (Opium); T40.1 (Heroin); T40.2 (Other opioids); T40.3 (Methadone); T40.4 (Other synthetic narcotics). The regression analysis was conducted using Joinpoint statistical software.

**Results:** The black population in the Midwest census region showed the highest age-adjusted mortality in the year 2020 (46.14 per 100,000). This was followed by the black (32.71 per 100,000) and white population (30.5 per 100,000) in the northeast census regions respectively. Overall, the opioid-related mortality followed a similar trend across all census regions. Except south census region where age-adjusted mortality was comparable between the black and white populations, blacks had higher opioid-related mortality in all other census regions.

**Conclusion:** This study provides concise evidence of inequality in opioid-related deaths among various US census regions. Policy changes focused to certain regions are required to significantly address the underlying factors related to disparities in opioid-related mortality among the black population.

## 1. Introduction

An estimate of 93,000 drug overdose deaths were reported in the United States in the year 2020(CDC, 2020). Among the various drugs reported, opioids were found to be the most commonly involved substance as a cause of mortality(Schiller, Goyal, & Mechanic, 2022). The opioid epidemic has been a public health concern in the US(Volkow & Blanco, 2021). Opioid-related mortality including prescription opioids, heroin and synthetic opioids has risen drastically since 1999(CDC, 2021). Availability and misuse of both prescription and illegal opioids, increased prescription of opioids, doctor shopping, increased use ofn heroine(Rosenblatt, Andrilla, Catlin, & Larson, 2015) have contributed to this opioid epidemic(McClellan et al., 2018). While programs such as the prescription drug monitoring program (PDMP)(Ali, Dowd, Classen, Mutter, & Novak, 2017) have shown a declining trend in obtaining illegal drugs, the overall national level opioid-related deaths continue to rise.

Several studies have described the trends of opioid-related mortality in the US over the last 20 years. There is evidence that the rising opioid epidemic is affecting disproportionately among various races, ethnicities, gender, and socio-demographic groups(Carpenter, McClellan, & Rees, 2017; Cicero, Ellis, Surratt, & Kurtz, 2014; Singhal, Tien, & Hsia, 2016). However, disparities in opioid-related mortality among states or census regions haven’t been explored yet. Understanding the inequalities and the trend of change in opioid-related mortality will help us make recommendations regarding the re-distribution of resources to alleviate crisis in certain regions. This study aims to provide insight into the difference in mortality between white and black population in various census regions of the country.

## 2. Methods

### 2.1. Data Collection and Measures

The data was extracted from the Center for Disease Control and Prevention Wide-ranging Online Data for Epidemiologic Research (CDC WONDER)(CDC, 1999-2020). This database includes county-level national mortality and population data from 1999 to 2020. The multiple cause of death files is compiled from data provided by the 57 vital statistics jurisdictions through the Vital Statistics Cooperative Program(CDC, 1999-2020). The data requested for this study included various International Classification of Disease (ICD-10) codes including F11.0 (Mental and behavioral disorders due to use of opioids, acute intoxication); F11.1 (Mental and behavioral disorders due to use of opioids, harmful use); F11.2 (Mental and behavioral disorders due to use of opioids, dependence syndrome); F11.3 (Mental and behavioral disorders due to use of opioids, withdrawal state); F11.4 (Mental and behavioral disorders due to use of opioids, withdrawal state with delirium); F11.5 (Mental and behavioral disorders due to use of opioids, psychotic disorder); F11.6 (Mental and behavioral disorders due to use of opioids, amnesic syndrome); F11.7 (Mental and behavioral disorders due to use of opioids, residual and late-onset psychotic disorder); F11.8 (Mental and behavioral disorders due to use of opioids, other mental and behavioral disorders); F11.9 (Mental and behavioral disorders due to use of opioids, unspecified mental and behavioral disorder); T40.0 (Opium); T40.1 (Heroin); T40.2 (Other opioids); T40.3 (Methadone); T40.4 (Other synthetic narcotics).

The extracted data from the CDC WONDER database included age-adjusted mortality rate (deaths per 100,000), 95% confidence intervals, and standard errors for White and Black non-Hispanic populations across various census regions of the United States. The US census regions of Northeast, Midwest, South, and West are based on the 2010 U.S. Census definitions(US_Census_Bureau, 2010).

### 2.2. Data Analysis

The data collected was stratified by year, race, and census regions as defined by the US Census Bureau. Due to a lack of data on other races, the analysis in this study included white and black populations only. The statistical analysis was conducted using Joinpoint regression statistical software. The use of Joinpoint software to study the trends in mortality rates over time is recommended for population-based death certificate data by the National Center for Health Statistics(Surveillance Research Program, 2020). The data downloaded on Microsoft Excel files were imported to the Joinpoint software program to run the regression analysis. The software used standard error data for calculating weighted least squares. Using time (in years) as the independent variable and age-adjusted mortality as the dependent variable, the software program executed regression analysis. The trends were reported as annual percent change (APC). It analyzed the continuous linear trends with changing points (time points), that is, joinpoints. This allowed us to test if a change in the trend is statistically significant.

## 3. Results

### 3.1 Northeast Census Region

There was a slight decrease in age-adjusted mortality rate (2.95 per 100,000 to 2.79 per 100,000) among the black non-Hispanic population between 1999-2011 (Figure 1). The annual percent change (APC) was calculated to be about −0.24% between this period (95% CI −3.3,2.9; p>0.05]. However, this population observed a significant increase in age-adjusted mortality rate (2.79 per 100,000 to 32.71 per 100,000) between 2011-2020 (APC 33.5%; 95%CI 30.4,36.6; p<0.0001). In comparison, the white population observed a significant increase in age-adjusted mortality rate throughout 1999-2020 (2.09 per 100,000 to 30.5 per 100,000). The annual percent change between 1999-2012 was 11.2% (95%CI 9.4-13; p<0.0001); 33.7% APC (95%CI 23.1,45.2; p-value<0.0001) between 2012-2016; and 7.3% APC (95%CI 3.6,11.2; p=0.0001) between 2016-2020. Overall, the black population saw an average APC of 13% (p<0.0001) as compared to 14.4% (p<0.001) in the white population between 1999-2020.

**Figure 1:**
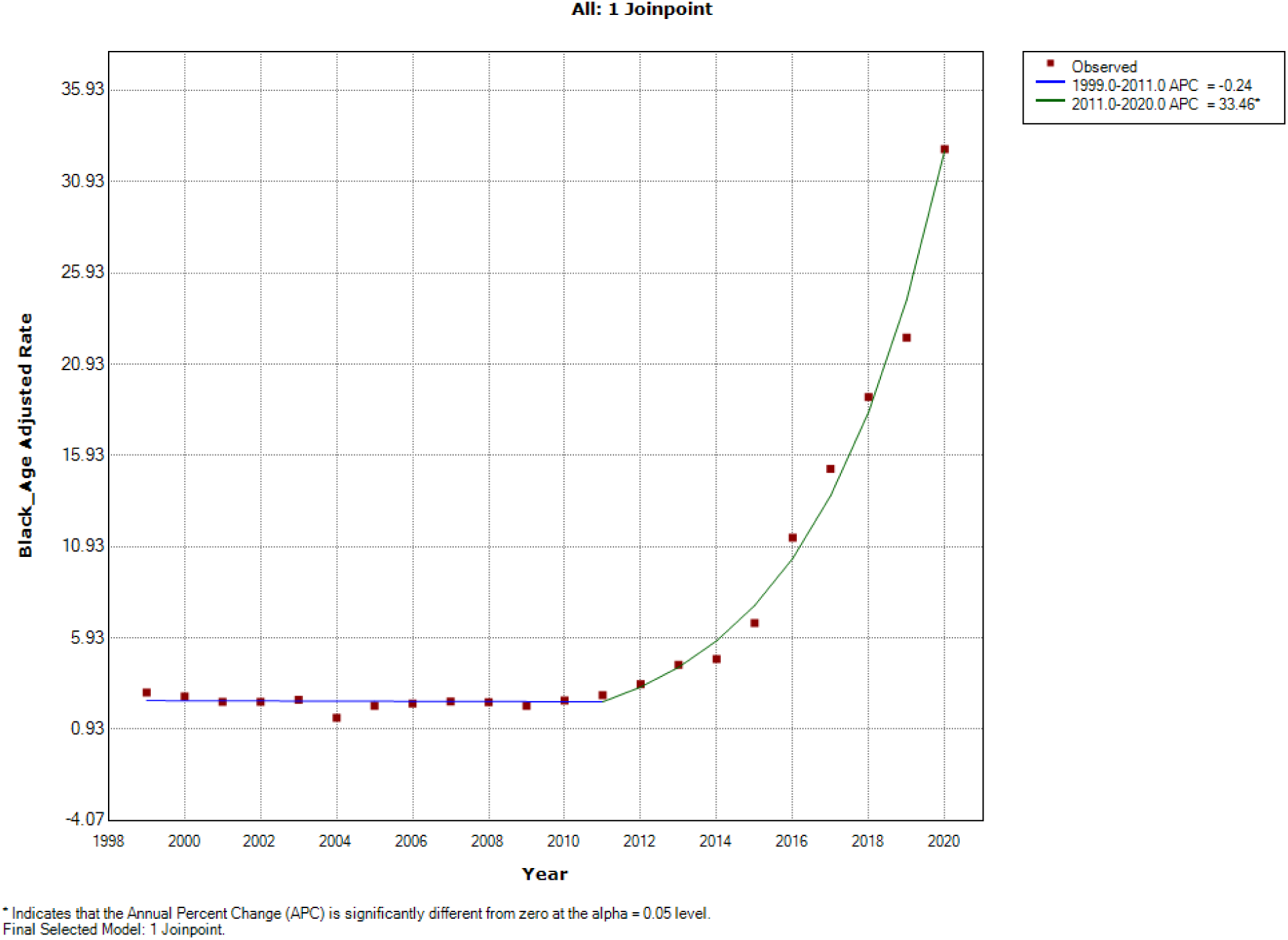
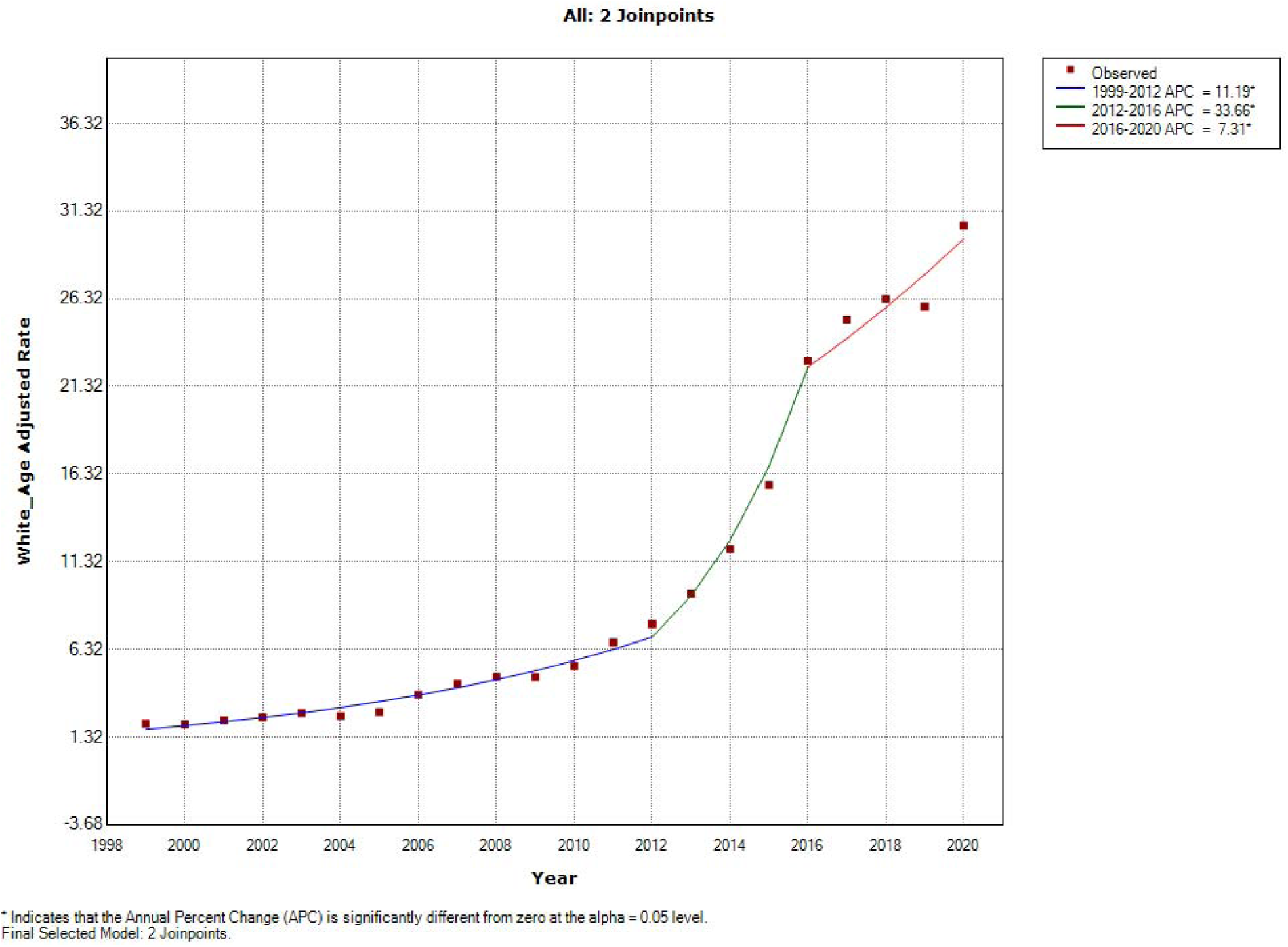
Northeast Census Region.

### 3.2 Midwest Census Region

During 1999 to 2011, the age-adjusted mortality rate increased from 2.52 per 100,000 to 5.75 per 100,000 among the black population. As shown in the figure 2, this represented a significant 5.3% APC (95% CI: 0.6,10.3; p=0.029). Similarly, there was a significant increase in mortality (5.75 per 100,000 to 46.14 per 100,000) in this population with an APC of 25.7% (95% CI: 21.8,29.8; p<0.001) between 2011-2020. On the contrary, the white population in the Midwest census region saw a steady increase in the age-adjusted mortality between 1999-2020 (0.98 per 100,000 to 22.84 per 100,000) with a significant APC of 14.6% (95%CI: 13.4,15.8; p<0.001). Overall, the black population saw an average APC of 13.6% (p<0.0001) as compared to 14.6% (p<0.1) in the white population between 1999-2020.

**Figure 2:**
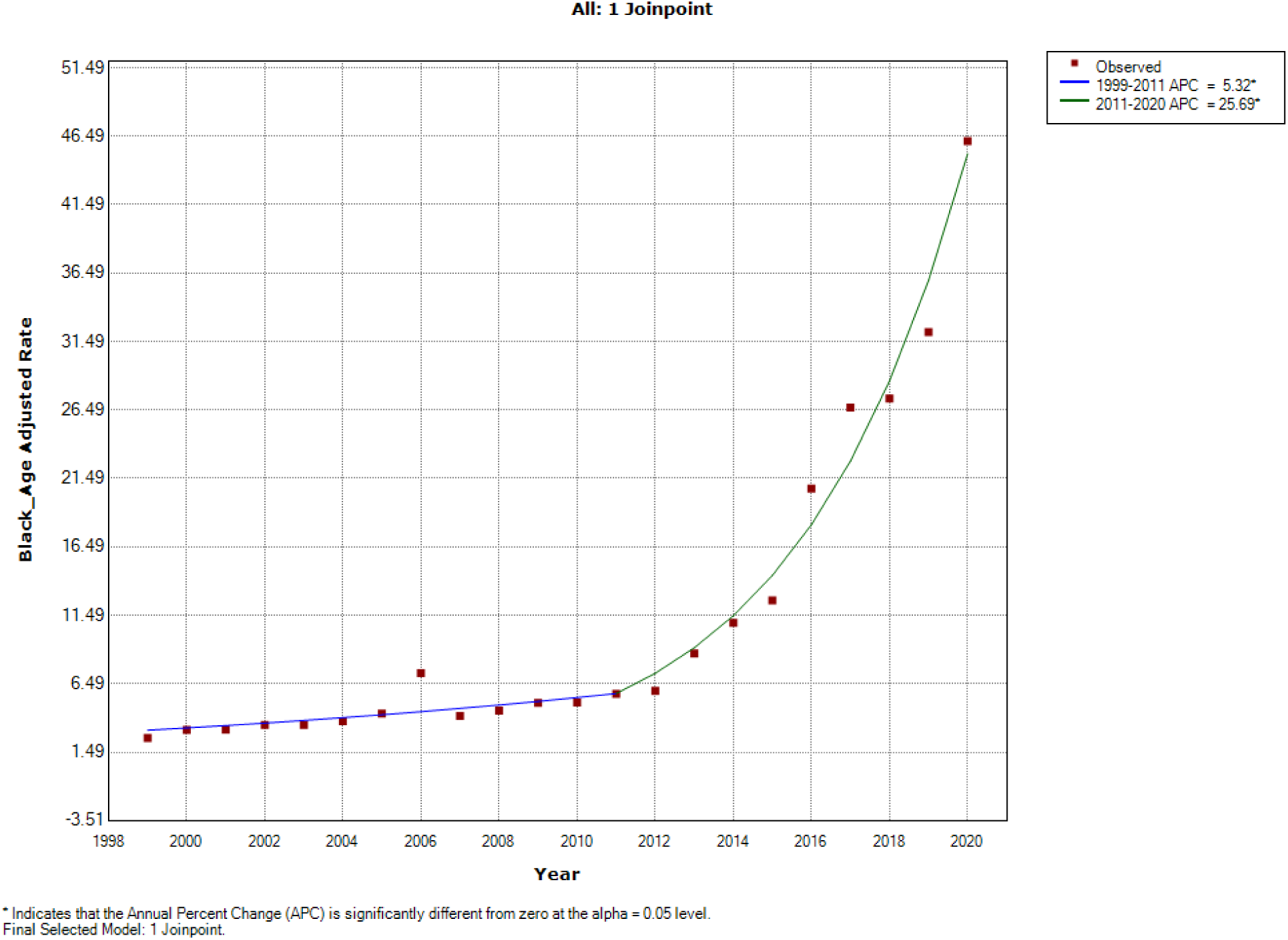
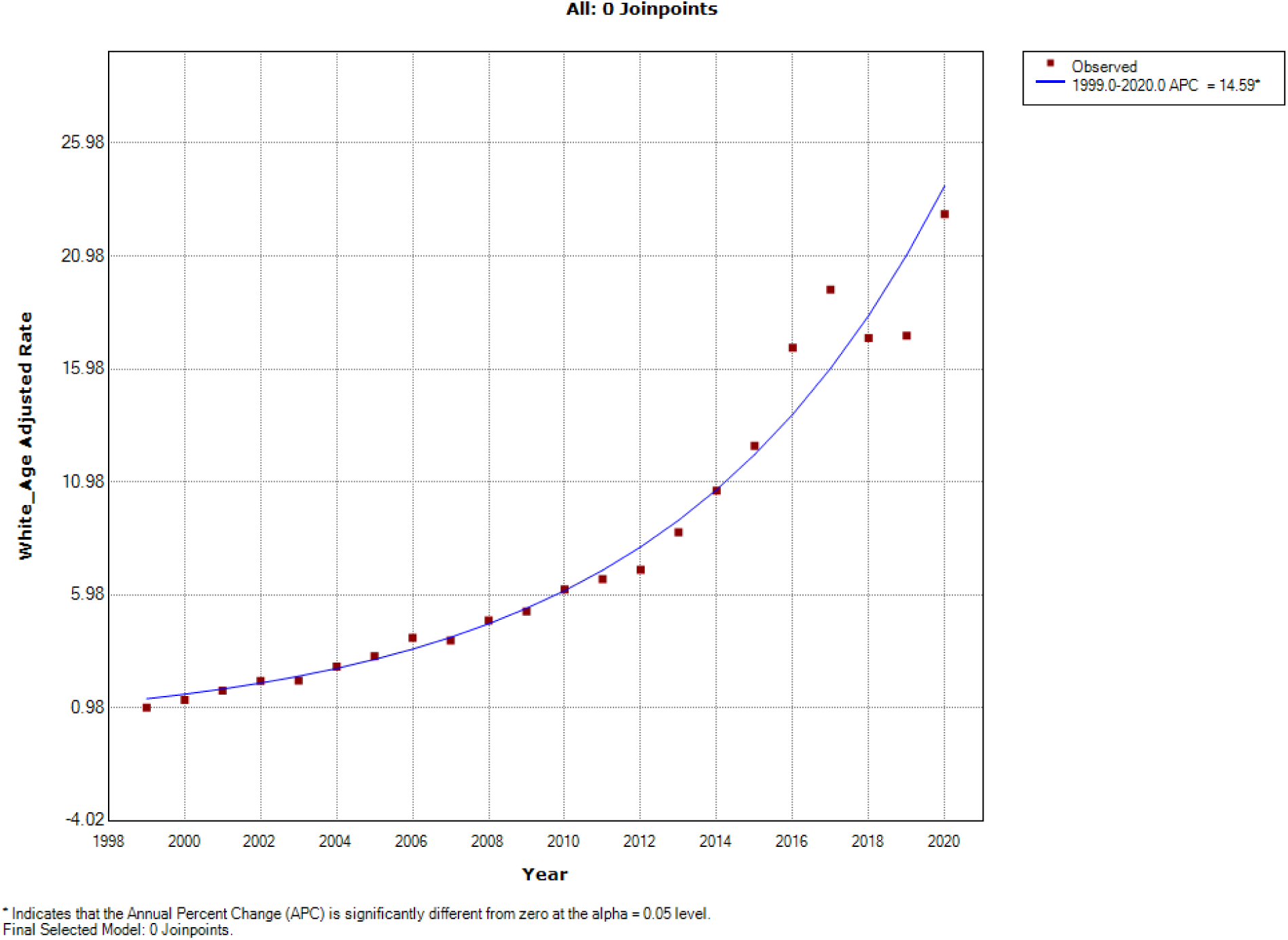
Midwest Census Region.

### 3.3 South Census Region

The age-adjusted mortality rate among black non-Hispanic population increased from 0.88 per 100,000 in 1999 to 18.96 per 100,000 in 2020 (Figure 3). The mortality changed at the rate of 6.3% APC (95%CI 2.8,10; p=0.001) between the time-period 1999-2012; APC was 29% (95%CI 25.3,32.7; p<0.001) between 2012-2020. Overall, the blacks saw an average APC of 14.5% (95% CI 12,17; p<0.001). On the other hand, the age-adjusted mortality in the white population steadily increased from 1.62 per 100,000 in 1999 to 23.95 per 100,000 in 2020. This represented an annual percent change in mortality of 11.1% (95% CI 10.1,12.1; p<0.001).

**Figure 3:**
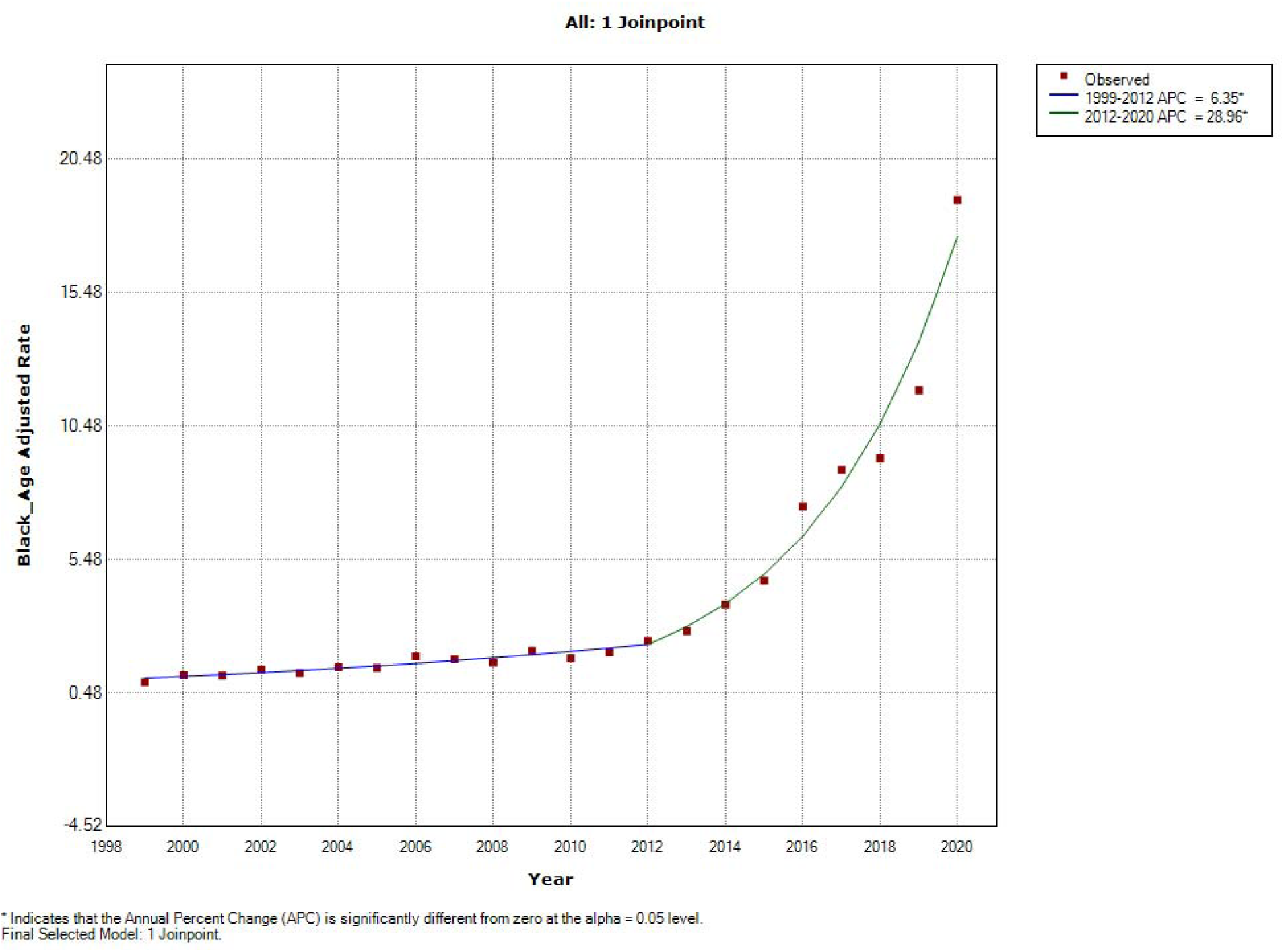
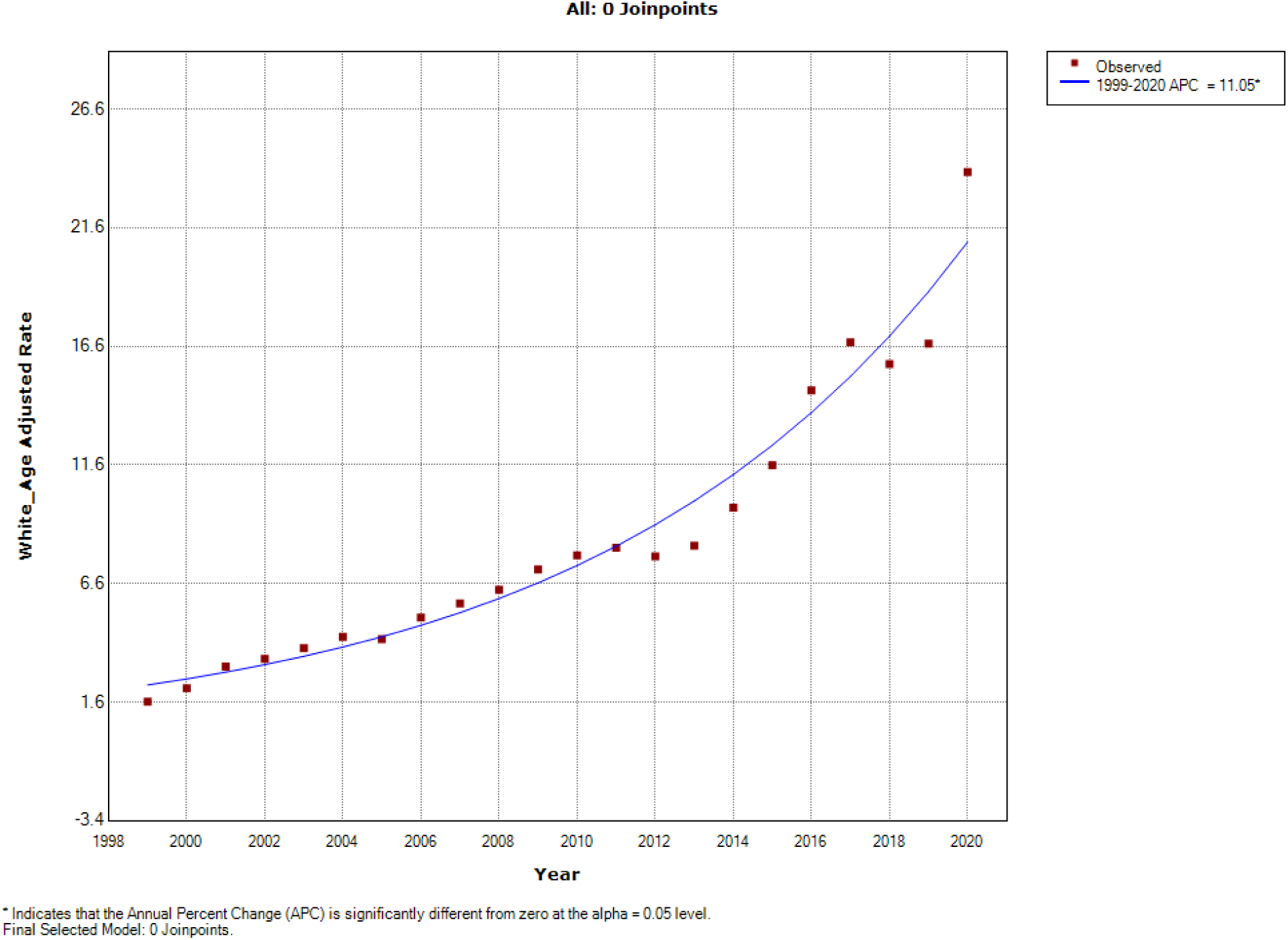
South Census Region.

### 3.4 West Census Region

The black population in the west census region saw a varied pattern of opioid-related mortality during 1999-2020. Figure 4 shows that there was a near-constant age-adjusted mortality between 1999-2012 (APC of 0.6%). However, the time period between 2012 to 2018 saw a significant increase in mortality by 10.7% APC (95% CI 4.5,17.3; p=0.002). This was followed by a sharper increase in age-adjusted mortality from 8.84 per 100,000 in 2018 to 22.35 per 100,000 in 2020. This represented an annual percent change of 63.6% (95% CI 40.5,90.7; p<0.001). Similarly, the white population saw a smaller but steady increase in opioid-related mortality until 2018 (APC 5%; p<0.001). But the annual change in mortality went up to 31.8% between 2018-2020 (95% CI 11.3,55.9; p=0.003). Overall, the average annual percent change in the age-adjusted mortality in the black population (8.3%; p<0.001) was higher compared to the white population (7.3%; p<0.1) in the west census region.

**Figure 4:**
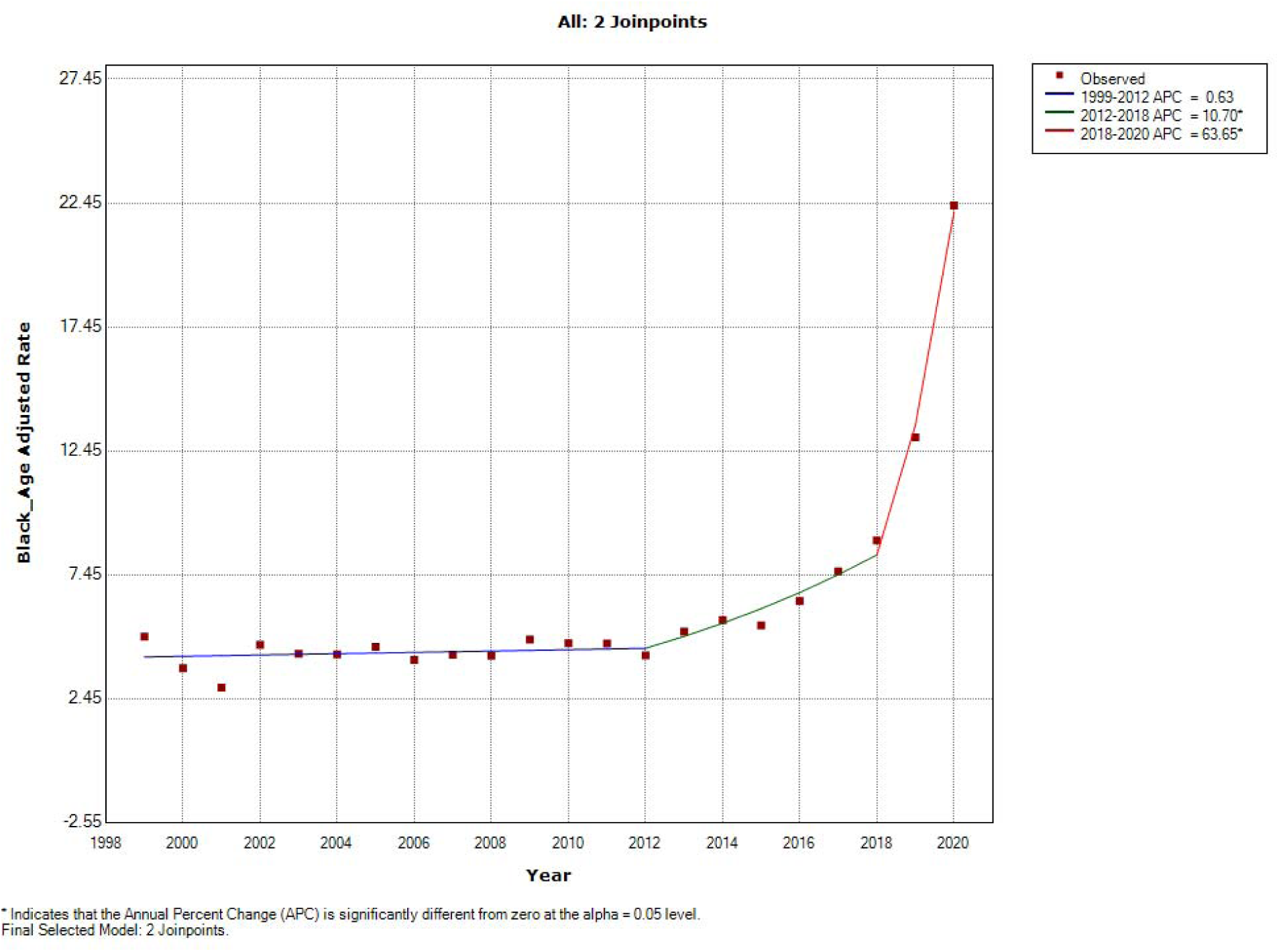
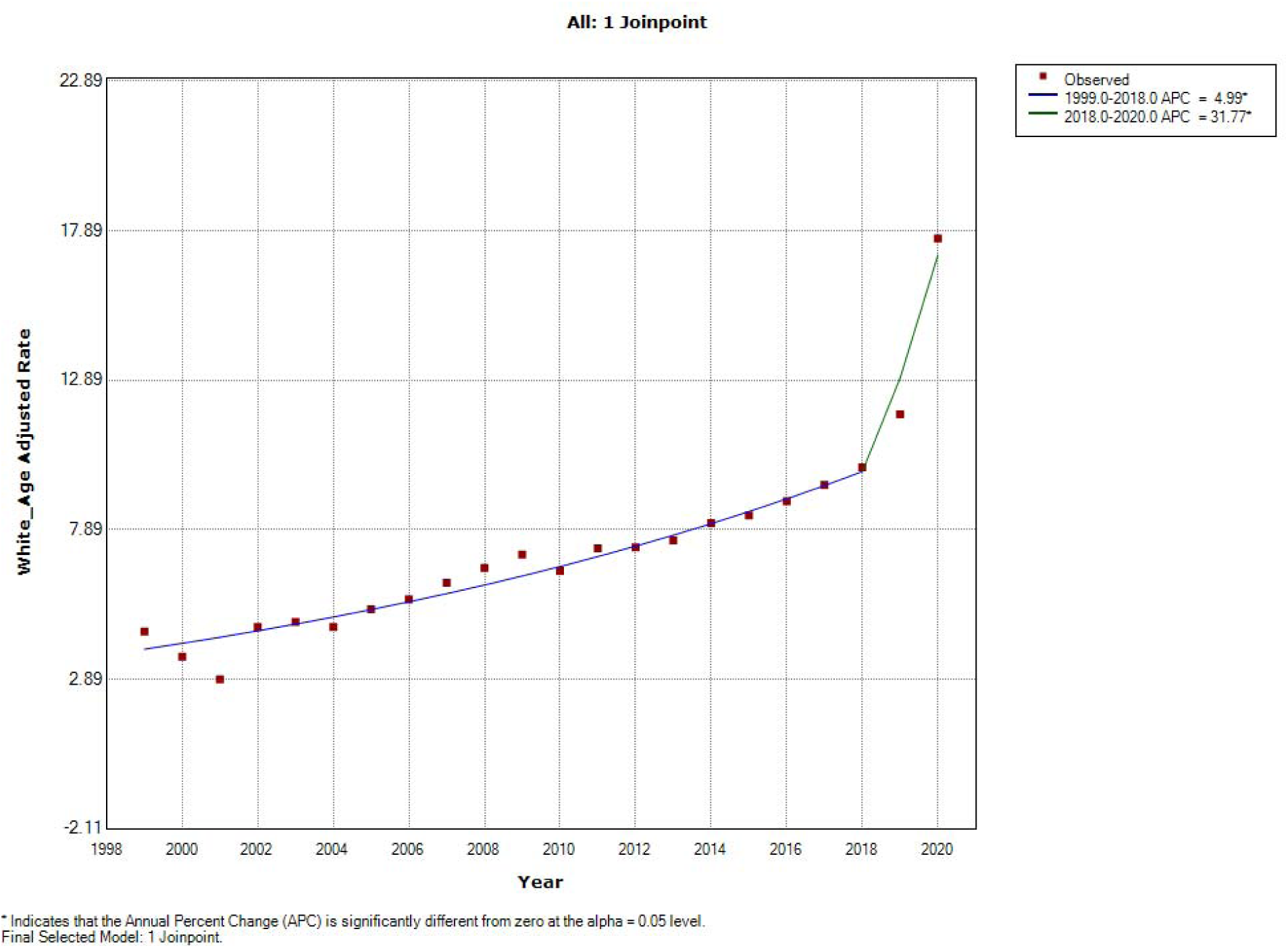
West Census Region.

## 4. Discussion

The statistical analysis conducted showed that the age-adjusted opioid-related mortality in the black and white populations has been consistently increasing since 1999. The blacks in the Midwest census region showed the highest age-adjusted mortality in the year 2020 (46.14 per 100,000). This was followed by the black (32.71 per 100,000) and white population (30.5 per 100,000) in the northeast census regions respectively. Overall, the trends in opioid-related mortality were found to be similar across various census regions. Except south census region where age-adjusted mortality was comparable between the black and white populations, blacks had higher opioid-related mortality in all other census regions.

Several studies have described disparities in opioid-related mortality due to race/ethnicity, education, and geographical distribution(Buchanich et al., 2016; Cerdá et al., 2017; Langabeer, Chambers, Cardenas-Turanzas, & Champagne-Langabeer, 2022; Stewart, Cao, Hsu, Artigiani, & Wish, 2017). The difference in socioeconomic status, in particular, unemployment and homelessness/housing insecurity, lack of education,(Svendsen, Fredheim, Romundstad, Borchgrevink, & Skurtveit, 2014) lack of health insurance has been shown to be contributing to this disparity(Saloner & B, 2013). Although the reasons were indistinct, opioid overdose was predominantly observed in men and women who were divorced, separated, or widowed. Behavioral, physical, and economic benefits complement having a marital relationship, reducing the risk of an individual predisposing to fatal opioid overdose. Developing interpersonal relationships is beneficial in building communities resistant to the opioid crisis and would help reduce opioid-related mortality(Altekruse, Cosgrove, Altekruse, Jenkins, & Blanco, 2020; Ellis, Kasper, & Cicero, 2020).

Altekruse et al. reported that adolescents, young, and middle-aged African American and White adults belonging to the low socioeconomic status were mainly affected with opioid fatality compared to those in the higher SES strata(Altekruse et al., 2020). The whites were shown to have relatively higher mortality rates due to opioid overdose, but this was overturned by black communities displaying higher opioid fatalities later during late 2000s. Similarly, the underprivileged minorities are more prone to occupational injuries leading to a higher risk of exposure to opioid analgesics or illicit drug use, resulting in fatal opioid overdose(Altekruse et al., 2020). Education and housing are known to be essential components of risky opioid use/overdose behavior. Individuals with lower levels of education are impacted more due to a lack of knowledge on drugs, partially related to unemployment, and lack of healthcare benefits might increase the risk of opioid relapse and overdose cases(Altekruse et al., 2020; Ellis et al., 2020). However, opioid use among adolescents and adults who seek opioids for improving their academic performance resulting in drug-seeking behaviors(Ellis et al., 2020).

Previous studies have shown that there is a disparity in treatment; blacks are less likely to receive treatment compared to non-Hispanic whites(Hollander, Chang, Douaihy, Hulsey, & Donohue, 2021). Structural differences in healthcare could be a contributing factor for racial disparities in opioid overdose. During the 1990s, the opioid crisis began in white communities through excessive prescribing of opioids compared to blacks(Altekruse et al., 2020). There were misperceptions and biases in the healthcare system, and blacks were undervalued for self-reporting of pain(Altekruse et al., 2020; SAMHSA, 2020). The negative stereotype behaviors of providers prompted them to seek illicit drug practices where the opioid analogs were readily available at low costs that led to drug overdoses in these communities(Schepis, Teter, & McCabe, 2018).

There has been extensive research comparing drug overdoses in urban vs rural areas(Altekruse et al., 2020; Ellis et al., 2020). Several studies have been able to identify counties and other regions where mortality is higher compared to other regions(Rossen, Khan, & Warner, 2013). This study has helped identify disparities in age-adjusted mortality rates among various US census regions. Understanding the pattern of change in mortality over different geographical regions can help address the opioid epidemic better. Harm reduction strategies or opioid treatment programs can be facilitated in regions where the epidemic is growing at a much higher rate. Targeted interventions(Hawk, Vaca, & D’Onofrio, 2015; McClellan et al., 2018) such as naloxone distribution or increased access to medication-assisted treatment might alleviate the sharp increase in opioid-related mortality and decrease the existing disparity.

### 4.1 Limitations

While CDC WONDER data gives us easy access to population health data, there could be several limitations as well. The data obtained from this database could be subjected to error. The data in this database is collected from death certificates which are subjected to inaccuracies in demographics or cause of death.

## 5. Conclusion

This study provides concise evidence of inequality in opioid-related deaths among various US census regions. State-wide policy changes or focused approaches are warranted in future to significantly address the underlying factors related to disparities in opioid-related mortality among the black population.

## Data Availability

All data produced in the present work are contained in the manuscript

